# Staff and Service User Experiences of Specialist Open Prison Personality Disorder Services: How do Pathways Enhanced Resettlement Services Support Progression?

**DOI:** 10.1101/2024.11.03.24316661

**Authors:** Georgina Mathlin, Hannah Jones, Carine Lewis, Claudia Cooper, Mark Freestone

**Author notes:** Corresponding Author: Georgina Mathlin.

## Abstract

**Background:** The Offender Personality Disorder (OPD) Pathway, a joint health and criminal justice initiative across England and Wales, aims to support rehabilitation of individuals with a likely diagnosis of personality disorder. Pathways Enhanced Resettlement Services (PERS) is an OPD service currently operating in five open prisons in England, which aims to support people at high risk of being returned to closed conditions or reoffending in the community after release. We aimed to understand service user and staff experiences of PERS.

**Methods:** We conducted semi-structured interviews with ten staff and nine service users. We then conducted a reflexive thematic analysis generating three themes.

**Results:** The three themes identified were: (1) “A shock to the system”, describing the challenges for service users posed by the liminal space of open prison (between higher security conditions and the community); and how in this context PERS might be viewed with suspicion but was for interviewees ultimately a space where they felt valued. (2) “We’ve got some understanding of their journey”; staff and service users described PERS staff developing more trusting relationships with service users than non-PERS staff, where service users felt understood and supported, practically and emotionally; and (3) “internal states can be real barriers to progression”; PERS staff supported service users to understand and overcome barriers, through enabling self-reflection, and tailoring support to times of greater stress, including key milestones such as parole boards or periods of trial leave.

**Conclusions:** Staff and service users feel PERS provides support to progress through open prison, through development of positive trusting relationships and individualised support in a challenging context where such support was not otherwise available.

## Background

The diagnosis of personality disorder is a complex mental health condition, characterised by poor relationship skills, issues with identity and, in some cases, a risk of physical harm to others (International Classification of Diseases version 11 (ICD-11); WHO, 2018). Personality disorder is a prominent issue in prison populations, with estimates of 65% of the prison population in western countries meeting the criteria (Fazel & Danesh, 2002). Personality disorder is associated with an increased risk of violence and recidivism (Coid et al., 2006; O’Driscoll et al., 2012; Duggan & Howard, 2009; Yu et al., 2012), substance misuse and self-harm (Blasco-Fontecilla et al., 2010; Yang et al., 2010). Working with individuals with a diagnosis of personality disorder has also been associated with high levels of staff burnout (Freestone et al., 2015).

The Offender Personality Disorder (OPD) Pathway is a joint health and criminal justice initiative in the United Kingdom commissioned by His Majesty’s Prison and Probation Service (HMPPS) and NHS England (Skett et al., 2017). The pathway was set up in 2012 to support high risk individuals with a likely diagnosis of personality disorder to mitigate some of the risk factors hindering successful rehabilitation in this group (Mathlin et al., 2024). It aims to reduce repeat serous offending, improve psychological wellbeing, produce a competent and trained workforce, and use resources efficiently. The OPD pathway is based on a whole system approach, recognising the stages of an offender’s journey with services in the community and prison. People are screened into the pathway using an algorithm (Jolliffe et al., 2012) and subsequently receive a formulation to inform sentence planning and suggest engagement with OPD services if appropriate.

In the UK, open prisons house “Category D” adult male offenders, who have a low risk of absconding and constitute a low risk to the public due to the nature of their offence, and those nearing the end of longer sentences who have transitioned from higher security categories (NOMS, 2015). The aim of open prisons is to aid the resettlement of prisoners back into society and provide opportunities for Release on Temporary License (ROTL). ROTL allow periods of short-term release to the community, to support prisoners to build links with family, housing services, volunteering and employment opportunities to facilitate their successful community integration on release (Auty & Liebling, 2020).

Open prisons allow greater freedom and autonomy, which can be difficult for those who have become institutionalised to closed condition prisons (Honeywell, 2015; Micklethwaite & Earle, 2021), often a factor for people screened onto the OPD pathway on long sentences. There are higher behaviour expectations of residents in open prisons and a different culture relative to closed conditions (Hacin & Meško, 2018). Retaining a position in an open prison can be difficult, due to the low threshold for being returned to a closed condition establishment, including substance misuse lapses, feeling threatened or bullied and being violent (Hallett & Lowbridge, 2014). The ease of being returned to closed conditions and witnessing people being returned creates an emotionally insecure environment in open prisons (Statham et al., 2020). Danks and Bradley (2018) found residents in open prisons were reluctant to seek support for mental health issues in case they were returned to a closed condition prison. Residents in open prisons are sometimes reluctant to use the complaint system, through fear of being labelled as problematic and removed (HMIP, 2018').

Five open prison OPD services known as Pathways Enhanced Resettlement Services (PERS) have been operational since 2019, focusing on offenders with a high risk of “failing” in the short-term, either through reoffending following release or being returned to closed conditions. Service users are identified through “red flags” which include being screened into the OPD pathway, having a high-risk of violent and/or general reoffending, an open plan to support risk of self-harm or suicide in the last 12 months and at least one prior failure in open conditions or the community within the 6 months prior to entering the open prions. PERS aim to support people through open conditions and for release into the community by reducing the chance of being returned to closed conditions and preparing for community transition.

Service users identified as suitable for the service join voluntarily and alongside living in the standard accommodation of the open prison, receive key work sessions where they develop a working formulation to anticipate difficulties and agree a management plan. In these key work sessions, service users are supported with community reintegration and developing social capital through release plans and accompanied Release on Temporary License (ROTL). The services have an open-door policy for individuals to ‘drop-in’ and receive *ad hoc* support and pre-empt crises.

Since the OPD pathway has been set up, numerous studies have explored the experiences of people using the variety of available OPD services. These studies consistently report the importance of developing trusting, supportive relationships with staff as a key mechanism for change (Bennett, 2014; Dolan, 2017; Duncan et al., 2022; Jarvis et al., 2022; King & Crisp, 2021; McMurran & Delight, 2017; Moran et al., 2022; Mullan et al., 2018; Shaw & Forster, 2018). Research with OPD staff has also been helpful to gain further insights into the functioning of the pathway. OPD staff have reflected on the personal growth and impact they can have working in an OPD service such as instilling hope and providing a safe environment (Akinsulire, 2020; Bond & Gemmell, 2014; Cooke et al., 2017; Moran et al., 2022).

Only one previous study has explored the experiences of PERS service users (Jarvis et al., 2022). It focussed on the initial pilot of the service in one open prison. Since then, PERS has evolved and been implemented across five open prison sites. It is therefore important to explore how PERS in their current form are experienced and whether they are supporting individuals to maintain their position in open prisons and progress into the community. The previous study focussed solely on service users but to provide a more holistic understanding of PERS the current study also focusses on staff. We therefore had two research questions:

1. How do staff and service users experience PERS?
2. Has the service supported individuals to remain in open prison and transition successfully to the community?

## Methods

### Ethical Approval

Ethical approval was granted by HMPPS National Research Committee (Ref: 2020-131/ Ref: 2021-180). All participants provided informed consent.

### Procedure and Sample

We conduced semi-structured interviews. We recruited PERS staff from the five services. We selected ten of twenty staff (two staff members from each service were approached), purposively, to ensure a variety of staff roles and levels of experience were recruited including both clinical and operational staff. Three groups of PERS service users were eligible to participate: ex-PERS service users in the community, current PERS service users and ex-PERS service users in closed conditions prisons. Potential participants were initially approached by PERS staff, directly or via offender managers of those returned to closed conditions. Staff then arranged interviews for current service users and passed details of ex-service users living in the community willing to participate to the researcher.

To determine an appropriate sample size, the information power framework was utilised (Malterud, Siersma & Guassora, 2016). The narrow study aim, dense sample specificity and expected high quality of dialogue within the interviews indicated a smaller sample would be adequate. However, as there is little extant research into PERS and the plan to analyse data at the group level suggested a larger sample would be most appropriate. Taking these criteria into consideration, a sample of 20 (ten staff and ten service users) was considered sufficient to address the research questions. The final sample size obtained was 19 (ten staff and 9 service users).

### Data Collection and Processing

Interviews with PERS staff were conducted by the lead author, remotely via Microsoft Teams in May 2021 whilst in-person research within prisons was prohibited due to COVID-19 restrictions. The co-authors developed a topic guide (S1), which explored respondents job role, and their views on how PERS supports users in open conditions; the interviewer probed potential barriers and facilitators of service users benefitting from PERS support. The topic guide also included questions regarding how COVID-19 impacted service delivery and service user outcomes.

Interviews with service users were conducted by the lead author in June 2022, with current service users interviewed in person at the open prisons and ex-service users interviewed remotely via telephone. We were unable to recruit any ex-service users in closed conditions, as no potential participants were identified. The topic guide (S2) developed for service user interviews asked how they had joined PERS, how PERS had been helpful or unhelpful, how COVID had impacted their use of PERS, what they feel they need to succeed, their wellbeing whilst working with PERS and, if applicable, their experience of transitioning into the community from PERS. All interviews were transcribed verbatim by the researcher who conducted the interviews.

### Data Analysis

NVivo Version 14 for Windows (QSR, 2020) was used to manage and organise the interview data. Reflexive Thematic Analysis (RTA; Braun & Clark, 2021) was applied to the data. Themes were generated from ideas that were covered widely in the interviews and/or important to the research question. Themes were reflective of the entire dataset, using an inductive approach where the analysis focused on the content of experiences reported, rather than exploring the language use to describe those experiences (Boyatzis, 1998; Trahan & Stewart, 2013). A summary of preliminary findings was shared via email with all PERS staff for comments and presented at a staff away day for discussion which aided the interpretation of the data.

### Researcher Reflexivity

The researchers have previously worked in OPD pathway services (although not PERS) and had prior knowledge and expectations about the experiences of staff and service users which likely influenced the interview questions and the interpretation of data. Prior experience of OPD services was a benefit during the interviews, as rapport was easily built with participants and due to prior understanding of the OPD context.

## Results

### Sample Characteristics

No demographic data was collected about gender, age or ethnicity of participants in line with the ethics agreement to maintain confidentiality in this sensitive area; however all service users were placed in male prisons. Ten staff were interviewed (two from each service). Staff were grouped as clinical (clinical leads and an assistant psychologist), and operational (mental health nurses and prison officers) based on their service role. Staff had worked in PERS between 3 months and 2 years (the clinical staff average was 13 months, and the operational staff average was 14 months). Three of the clinical staff and three of the operational staff had previously worked in other OPD Pathway services.

Nine service users were interviewed. Seven were current PERS service users and two were ex-PERS users living in the community. The service users had been in open prison between 5 months and 3 years. All service users had been in higher category prisons prior to entering open prison, two of the current service users had been in open prison previously (and been returned to closed conditions) and two service users had been recalled from the community during their sentence.

### Thematic analysis

We developed three themes that encompass the experiences of PERS reported by staff and service users and provide insights into the functioning of PERS and how service use supports progression through open prison.

#### *Theme 1: “*A shock to the system”: managing the liminal space of open prison

Service users described adapting to open prison as “*hard work” [SU4]*, “*difficult” [SU7]* and “*strange” [SU5].* Service users felt the processes in open prison were confusing and experienced “*not knowing where to seek help, how to ask for help here” [SU5].* The different routine of open prison compared to closed conditions, especially the increased freedom, was described as “*a shock to the system” [SU2]*. Alongside adapting to the freedom of open prison, service users expressed fear that this freedom could easily be removed by the general open prison staff. For example, one participant described: “*you say the wrong thing to a member of staff, you could get a nicking and then you’re going to get a ROTL ban” [SU4].* PERS staff commented on the greater freedom of choice for service users in the open prison setting too, including choices that would pose risks to progression. For example, “*it’s really easy for them to get drugs if they want” [OS1]* and “[there’s] *less staff, less security, less everything to stop that” [OS1]*.

The initial motivation for some service users to join PERS was because they were “*searching for that person to talk to, to understand how I was feeling… I wasn’t getting that via the units” [SU5].* Service users reflected wanting someone to talk to about the difficulties of being in open prison alongside getting support with issues such as self-harm, personality disorder and progression. This support was not felt to be available in the wider open prison.

Despite many service users having a desire to join PERS to gain support, some service users expressed concerns at how engaging with it might affect their progression through the system. For example, one participant described: “*I was a bit sceptical, thinking hold on a minute, does this mean I’m going back on the bus and I’m going back to closed conditions” [SU6].* There was suspicion about what PERS was, with one participant feeling that “*it’s a set up” [SU4]*. Staff reflected on the initial wariness some service users experienced when approached to join PERS, stating the importance of encouraging voluntary engagement: “*the service is voluntary; it therefore is quite non-threatening which enables people to engage with us” [CS4].* When service users were given more information about PERS and told that they had a choice to engage, they felt more open to joining: “*the more I was chatting with them, I was thinking yeah you know what, they’re actually not bad” [SU4]*.

PERS staff explained the range of support available through PERS. The clinical staff described “*it’s that emotional containment that feels important*” *[CS2]*, which they felt was developed through consistent key work and having drop-ins where service users receive ad-hoc support. Service users were positive about being able to use drop-ins “*you can just knock the door and walk in, and anyone of them will be happy to give you some advice and encourage you” [SU2].* Operational staff focussed more on the practical support they provide, for example “*a lot of the guys don’t know how to use a mobile phone, we take that individual on a day out, we’ll go through it…, there’s no panic” [OS1].* Service users reflected positively on the support available through PERS, and how they felt valued by it. Group activities such as tea and coffee evenings “*made me feel normal” [SU5]*. There was a clear emphasis that staff have time to provide this support to service users “*even if they’ve got a busy day, they’ll take the time out” [SU4]*.

Service users accessing PERS valued having “*somebody to talk to about how I was adapting” [SU9]* and being able to discuss concerns about freedoms being removed with PERS staff was helpful: “*coming here and letting that [concerns of being removed] off to them, I find it real helpful” [SU5].* Staff highlighted “*being really, really realistic with people… helping them structure expectations” [CS4]* was important when service users first enter open prison and that managing service user expectations of open prison helped service users adapt.

#### Theme 2: “We’ve got some understanding of their journey” engaging differently with service users

Service users reported having positive, trusting relationships with PERS staff where they felt “*relaxed around them, more open, honest” [SU6].* PERS staff were described as “*friendly and helpful” [SU1]* and that “*they listen to what you have to say” [SU8].* Service users described how they used PERS to offload worries “*I come up, I vent, I vent, I vent, and then I leave here and it’s just like, done’ [SU5].* Staff felt building relationships with service users was important for progression whilst also reflecting on how difficult it can be for the service users to develop trusting relationships “*a lot of them don’t trust” [OS6]*. They spoke about how they developed the trusting relationships citing “*giving people praise” [OS6]* and giving them the space to offload their worries as useful techniques to develop trust.

Service users described PERS staff as being available: “*I can come over here any time of day” [SU4]* which was different to how they experienced non-PERS staff in open prison, who were described as “*not very helpful” [SU7].* For example, one participant described how they often feel dismissed when approaching standard wing officers to have a chat “*[they say] ‘you’ll be alright mate, don’t worry about it’ and close you behind your door when in reality, you just need a chat for ten minutes” [SU6].* Service users also felt misunderstood by non-PERS staff “[*the wing officers were*] *looking at me like I’m speaking a foreign language” [SU7].* PERS staff recognised that the relationships service users have with them were better than non-PERS staff: “*We develop a far better…relationship with them than…the wing staff, sadly” [OS3]*. PERS staff feel they are “*more geared with emotional support” [OS4]* and have “*more time to help” [OS4].* Staff also felt they had a better understanding of service users than the general prison staff “*we’ve engaged with them in a different way, we’ve got some understanding of their journey…that in general the prison staff don’t have” [CS2]*.

#### Theme 3: “Internal states can be real barriers to progression” understanding and overcoming barriers

PERS staff felt that the “*internal states can be real barriers to progression” [CS1]* such as “*anxiety”* and when individuals have become “*hopeless and disempowered” [CS4]* which may have developed due to having a limited sense of control. Service users reflected on how they felt they were able to overcome some of the internal barriers to progression by working with PERS. For example, one participant described how he “*used to class myself as scum” [SU5]* and felt no matter what was said, he had always previously felt like this. Some other service users described how “*most of my goals and my targets haven’t really been realistic or achievable” [SU7]* and that they use to be “*angry and I’d take my frustration out on staff or another prisoner” [SU5]*. However, through working with PERS, service users felt they had been able to move past some of these difficult internal states. For example, “*one of the biggest things for me, was never forgiving myself” [SU5].* Service users felt they were “*a lot more aware of how my actions affect others”* and described being able to “*set realistic targets” [SU7]* and as a consequence service users experienced being “*a lot […] happier” [SU5]* following their engagement with PERS.

Service users felt they were supported to progress through their sentence, especially around key events such as upcoming parole boards. For example, one participant reflected how their contact with PERS increased as they approached their parole board, to get support with their anxiety: “*I see them every week now, because of the anxiety” [SU5].* Overall service users felt PERS “*try to move people forward through the system” [SU1].* However, progression during COVID-19 was impacted, as “*people were not getting parole because they hadn’t been able to access ROTL, essentially what they’ve come to open conditions to do” [CS4]*. Some staff commented that “*[service users] got released when they’d had just had a couple of ROTLs, which never user to happen [before COVID]” [OS6].* Although it was positive to see that some people were able to progress in this time, there was also concern that individuals released without utilising ROTL may have missed out on opportunities to “*gain qualifications”* and “*bolster their support networks” [OS5]*.

### Reflective Statement

Staff and service users were willing and open to participate in the study and explore their experiences of PERS and how they felt PERS influences progression. Staff generally had similar views about their experiences of implementing PERS and the factors that influence progression, however, clinical staff tended to focus on the wider systems that service users exist within and the emotional difficulties they experience, whilst operational staff provided more detail on the day-to-day service delivery and the practical support they provide. This is likely to reflect their roles and the nature of their contacts with service users. Some service users expressed their dislike toward authority and directly referenced the interviewer being an authority figure that previously they would not have felt comfortable speaking with. They felt their work with PERS allowed them to be more open with authority figures and therefore take part in these interviews.

## Discussion and conclusion

This is the first study that explored the experiences of staff and service users across all five PERS services. Ten PERS staff and nine current and ex-service users took part. Their responses provide an insight into how staff and service users experience PERS and how they feel engaging with the service can support progression.

Many of the service users’ motivation to engage with PERS was to gain support with the transition into open prison alongside issues such as self-harm and their personality disorder diagnosis. This was not a universal experience however, and some service users initially felt suspicious about what PERS was. The reluctance to engage with PERS supports research exploring factors which may impact the readiness to engage in services. The suspicion and reluctance to engage in PERS highlights potential negative perceptions of the service, which is likely linked to a lack of trust in others (McMurran, 2012). PERS staff were aware of the reluctance some service users expressed about joining the service but reduced barriers to engaging by providing clear information about the service and a providing a choice about whether to engage. This in turn allowed service users to start to develop trust with PERS staff.

Moving to open prison was described as a difficult transition by all service users, where many struggled to adapt to the increased amount of freedom. This finding is supported by research, where people reported feeling overwhelmed when they first came into the open prison (Statham et al., 2021). PERS staff were aware of how difficult this transition could be and reported managing expectations as a useful tool to help service users adapt to their new environment. Expectations of treatment have been explored as a factor linked to treatment non-completion within offenders with personality disorder (McMurran, 2012) but have not been considered in relation to standard prison transitions.

Service users and staff highlighted the positive trusting relationships they experienced working with PERS. Building trusting relationships has been a consistent theme related to change in other OPD pathway services (Craissati et al., 2021; McMurran & Delight, 2017) and in a previous study exploring a pilot PERS service (Jarvis et al., 2022). Trusting relationships were seen as a key mechanism for progression within PERS. Trusting relationships increased initial engagement with the service and also enabled emotional and practical support to be provided in a meaningful way.

Contrastingly, many staff and service users spoke about poor relationships with non-PERS staff in the open prisons such as feeling misunderstood by non-PERS wing officers. Poor relationships have been reported in research exploring open prison environments (Danks & Bradley, 2018; Micklethwaite & Earle, 2021; Statham et al., 2021) due to residents feeling staff have additional powers to remove the privileges of being in an open prison. Poor relationships with staff in open prisons are concerning, especially for those with a likely diagnosis of personality disorder, as this group already have an increased likelihood of issues with interpersonal functioning and authority (Farnam & Zamanlu, 2018). An intended consequence of PERS is to improve all service user relationships however the experiences of poor relationships between service users and non-PERS staff suggests there is room for improved relationships with non-OPD professionals. Furthermore, a broad aim of the OPD pathway is workforce development (Skett & Lewis, 2019) with the added benefit of systemic change in the wider environments that OPD services operate within. PERS staff should therefore focus further on providing supervision and support to prison staff in open prisons to help develop better relationships with service users.

PERS was felt to be supporting progression through the open prison and preparing people for release into the community. However, the impact of COVID-19 was most felt in relation to progression, either due to not being granted parole boards or getting parole without any of the support such as gaining qualifications and increasing social support networks prior to release. Currently little research exists exploring the impact of COVID-19 on progression and re-entry to the community in the UK. The number of prison releases following the COVID-19 pandemic (March 2020) reduced by 15% compared to the same period in 2019 (Offender Management Statistics Quarterly, 2020) but this may be attributable to the prison population serving longer sentences, rather than a direct effect of COVID.

Aside from COVID-19, several barriers to progression were noted by staff and service users. The accessibility of drugs in the open prison estate and the reduced levels of security to monitor this were noted by staff. Low levels of hope and control were also considered as barriers to progression. There are associations between externalising control and offending behaviour (Turkcapar et al., 2002), with greater internal control associated with hopefulness (Mutlu et al., 2010). Having hope and a sense of internal control are likely to aid people through the difficulties of progressing in open prison and into the community (Craissati et al., 2021). Increasing levels of hope and control is therefore an important task of PERS and was seen through many service users reporting increased wellbeing, a newfound ability to forgive themselves and the ability to set more realistic goals.

Overall, PERS service users reported benefitting from the quick contact made by PERS following entry to the open prison, as well as ongoing support towards progression and release. The whole journey support indicates PERS is modelling the OPD pathway approach well, which intends to provide support across the various stages of a person’s journey through the CJS (Joseph & Benefield, 2012). This is especially important in relation to the context of open prison, an important stage of progression in many offender journeys that can be difficult to manage (Micklethwaite & Earle, 2021).

## Limitations

A cross-section of operational and clinical PERS staff were recruited, ensuring a broad range of staff perspectives were included. PERS staff may have been biased in reporting the positive impact of PERS however, as they have an interest in the continuation of the service. Likewise, the sampling of service users through the PERS staff may have meant only those who felt positively towards the service were approached to take part. Despite this, the consistent message of trusting relationships being developed in PERS and the feeling of whole journey support through the open prison indicates this is likely a reality for many PERS service users. It was not possible to recruit ex-service users returned to closed conditions in the current study and it will be important to understand whether this group feels similarly.

## Future Research, Clinical and Policy Implications

Future research into PERS should further explore the experiences of ex-service users who have been returned to closed. An understanding of how and why PERS was not able to support this group through open prison was not developed in the current study but is important to fully understand the functioning of PERS and how it can support progression. Furthermore, long-term outcomes (such as reoffending and recall, housing and employment when released into the community) of PERS service users should be reported on, monitored, and compared to non-PERS comparison groups where possible.

The findings of this study provide a useful insight into how PERS functions and supports progression. PERS should continue to provide individualised support for service users, addressing needs on a person-by-person basis. The relationships developed between staff and service users should continue to be a key focus alongside provision of key work, informal events, accompanied ROTL and drop-ins. Further consideration should be given to the relationships developed between service users and standard open prison staff, with PERS staff working closely with the open prison to increase the understanding of and ways of working with the PERS population.

As part of the new UK government’s focus on reforming the prison system, services such as PERS are vital. The Chief Inspector of Prisons has highlighted that resettlement support is “stretched” and it has been highlighted by charities that support for help with mental health is inadequate. This study shows PERS provides enhanced emotional and practical support to aid resettlement and provides a framework for how resettlement and mental health support could be improved more widely across the prison estate.

## Data Availability

Due to the sensitive nature of the work and for confidentiality purposes, the ethics committee did not permit making data produced in the study available.

## Declarations

## Acknowledgments

We thank the staff and service users who took part in this study and provided feedback on the findings.

## Author contributions

All authors read and approved the final manuscript.

## Availability of data and materials

Interview transcripts are not available in line with the ethics agreement for this research.

## List of Abbreviations

OPD: Offender Personality Disorder
PERS: Pathways Enhanced Resettlement Service
ROTL: Release on Temporary Licence

## References

Akinsulire, O. (2020). Exploring the experience of emotional stress and burnout in nursing health care professionals working in personality disorder forensic settings. City, University of London.

Auty, K. M., & Liebling, A. (2020). Exploring the relationship between prison social climate and reoffending. Justice Quarterly, 37(2), 358–381.

Bennett, A. L. (2014). Service users’ initial hopes, expectations and experiences of a high security psychologically informed planned environment (PIPE). Journal of Forensic Practice.

Bond, N., & Gemmell, L. (2014). Experiences of prison officers on a lifer psychologically informed planned environment. Therapeutic Communities: The International Journal of Therapeutic Communities.

Boyatzis, R. E. (1998). Transforming qualitative information: Thematic analysis and code development. sage.

Braun, V., & Clarke, V. (2021). Can I use TA? Should I use TA? Should I not use TA? Comparing reflexive thematic analysis and other pattern-based qualitative analytic approaches. Counselling and Psychotherapy Research, 21(1), 37–47.

Blasco-Fontecilla, H., Baca-Garcia, E., Duberstein, P., Perez-Rodriguez, M. M., Dervic, K., Saiz-Ruiz, J., Courtet, P., De Leon, J., & Oquendo, M. A. (2010). An exploratory study of the relationship between diverse life events and specific personality disorders in a sample of suicide attempters. Journal of Personality Disorders, 24(6), 773–784.

Coid, J., Yang, M., Tyrer, P., Roberts, A., & Ullrich, S. (2006). Prevalence and correlates of personality disorder in Great Britain. The British Journal of Psychiatry, 188(5), 423–431

Cooke, E., Stephenson, Z., & Rose, J. (2017). How do professionals experience working with offenders diagnosed with personality disorder within a prison environment? The Journal of Forensic Psychiatry & Psychology, 28(6), 841–862.

Craissati, J., Ramsden, J., Ryan, S., Webster, N., & West, L. (2021). Intensive intervention and risk management services (IIRMS) three years on: what we need to do better in the offender personality disorder pathway. The Journal of Forensic Practice

Danks, K., & Bradley, A. (2018). Negotiating barriers: prisoner and staff perspectives on mental wellbeing in the open prison setting. Journal of Criminal Psychology.

Dolan, R. (2017). HMP Grendon therapeutic community: the residents’ perspective of the process of change. Therapeutic Communities: The International Journal of Therapeutic Communities.

Duggan, C., & Howard, R. (2009). The ‘functional link’ between personality disorder and violence: A critical appraisal. Personality, personality disorder and violence, 19–37.

Duncan, K., Winder, B., Blagden, N., & Norman, C. (2022). “I’ve got the energy to change, but I haven’t got the energy for this Kinda therapy”: A qualitative analysis of the motivations behind democratic therapeutic community drop-out for men with sexual convictions. International journal of offender therapy and comparative criminology, 66(12), 1213–1236. p

Farnam, A., & Zamanlu, M. (2018). Personality disorders: The reformed classification in international classification of Diseases-11 (ICD-11). Indian Journal of Social Psychiatry, 34(5), 49.

Fazel, S., & Danesh, J. (2002). Serious mental disorder in 23 000 prisoners: a systematic review of 62 surveys. The Lancet, 359(9306), 545–550.

Freestone, M. C., Wilson, K., Jones, R., Mikton, C., Milsom, S., Sonigra, K., Taylor, C., & Campbell, C. (2015). The impact on staff of working with personality disordered offenders: A systematic review. PloS one, 10(8), e0136378.

Hacin, R., & Meško, G. (2018). Prisoners’ perception of legitimacy of the prison staff: A qualitative study in Slovene prisons. International journal of offender therapy and comparative criminology, 62(13), 4332–4350.

Hallett, E., & Lowbridge, C. (2014). Why do prisoners take the risk. BBC News,[online] http://www.bbc.co.uk/news/uk-england-27292555

Honeywell, D. (2015). Doing time with lifers: A reflective study of life sentence prisoners. British Journal of Community Justice, 13(1), 93–104.

HM Inspectorate of Prisons (2018). Report on an unannounced inspection of HMP Kirkham

Jarvis, D., Shaw, J., & Lovell, T. (2022). Service user experiences of a psychologically enhanced resettlement service [PERS] in an English open prison. The Journal of Forensic Practice(ahead-of-print).

Jolliffe, D., Cattell, J., Raza, A., & Minoudis, P. (2017). Factors associated with progression in the London pathway project. Criminal behaviour and mental health, 27(3), 222–237.

Joseph, N., & Benefield, N. (2012). A joint offender personality disorder pathway strategy: An outline summary. Criminal Behaviour and Mental Health, 22(3), 210–217.

King, N., & Crisp, B. (2021). Conceptualising ‘success’ among Imprisonment for Public Protection (IPP) sentenced offenders with personality-related difficulties. Probation Journal, 68(1), 85–106.

Malterud, K., Siersma, V. D., & Guassora, A. D. (2016). Sample size in qualitative interview studies: guided by information power. Qualitative health research, 26(13), 1753–1760.

Mathlin, G., Freestone, M. & Jones, H. (2024) Factors associated with successful reintegration for male offenders: a systematic narrative review with implicit causal model. Journal Experimental Criminology 20, 541–580. 10.1007/s11292-022-09547-5

McMurran, M. (2012). Readiness to engage in treatments for personality disorder. International Journal of Forensic Mental Health, 11(4), 289–298.

McMurran, M., & Delight, S. (2017). Processes of change in an offender personality disorder pathway prison progression unit. Criminal Behaviour and Mental Health, 27(3), 254–264.

Micklethwaite, D., & Earle, R. (2021). A Voice Within: An Autoethnographic Account of Moving from Closed to Open Prison Conditions by a LifelJSentenced Prisoner. The Howard Journal of Crime and Justice, 60(4), 529–545.

Moran, P., Jarrett, M., Vamvakas, G., Roberts, S., Barrett, B., Campbell, C., Khondoker, M., Trebilcock, J., Weaver, T., Walker, J., Crawford, M., & Forrester, A. (2022). National Evaluation of the Male Offender Personality Disorder Pathway.

Mullan, C., Johnson, D., & Tomlinson, J. (2018). Personality disordered offenders’ experiences of completing social skills treatment. *Journal of Criminological Research*, Policy and Practice.

Mutlu, T., Balbag, Z., & Cemrek, F. (2010). The role of self-esteem, locus of control and big five personality traits in predicting hopelessness. Procedia-Social and Behavioral Sciences, 9, 1788–1792.

O’Driscoll, C., Larney, S., Indig, D., & Basson, J. (2012). The impact of personality disorders, substance use and other mental illness on re-offending. Journal of Forensic Psychiatry & Psychology, 23(3), 382–391.

Offender Management Statistics Quarterly. (2020, April to June). Retrieved from https://assets.publishing.service.gov.uk/media/5f99adc18fa8f57f34060d26/Offender_Management_Statistics_Quarterly_April_to_June_2020.pdf

QSR International Pty Ltd. (2020) NVivo (released in March 2020) https://www.qsrinternational.com/nvivo-qualitative-data-analysis-software/home

Skett, S., Goode, I., & Barton, S. (2017). A joint NHS and NOMS offender personality disorder pathway strategy: A perspective from 5 years of operation. In: HeinOnline.

Skett, S., & Lewis, C. (2019). Development of the Offender Personality Disorder Pathway: A summary of the underpinning evidence. Probation Journal, 66(2), 167–180.

Statham, B. M., Winder, B., & Micklethwaite, D. (2021). Success within a UK open prison and surviving the ‘pains of freedom’. Psychology, Crime & Law, 27(8), 729–750.

Trahan, A., & Stewart, D. M. (2013). Toward a pragmatic framework for mixed-methods research in criminal justice and criminology. Applied Psychology in Criminal Justice, 9(1).

Turkcapar, M. H., Ozel, A., Guriz, O., & Isik, B. (2002). Locus of control in antisocial personality disorder. Cognitive Psychotherapy Toward a New Millennium: Scientific Foundations and Clinical Practice, 381-384.

Turley, C., Payne, C., & Webster, S. (2013). Enabling features of Psychologically Informed Planned Environments. London: Ministry of Justice analytical series. https://assets.publishing.service.gov.uk/government/uploads/system/uploads/attachment_data/file/211730/enabling-pipe-research-report.pdf

WHO. (2018). ICD-11 - Mortality and Morbidity Statistics

Yang, M., Coid, J., & Tyrer, P. (2010). Personality pathology recorded by severity: national survey. The British Journal of Psychiatry, 197(3), 193–199.

Yu, R., Geddes, J. R., & Fazel, S. (2012). Personality disorders, violence, and antisocial behavior: a systematic review and meta-regression analysis. Journal of Personality Disorders, 26(5), 775–792.

