## Supplementary material for "Staff and Service User Experiences of Specialist Open Prison Personality Disorder Services: How do Pathways Enhanced Resettlement Services Support Progression?": S1, S2

**Supplement 1:**

**Staff Interview Schedule:**

| Topics | Main Questions |  | Follow-up Questions | | | |
| --- | --- | --- | --- | --- | --- | --- |
| 1) Introduction of myself/ the topic/ boundaries of the interview/ consent. |  |  |  | | | |
| 2) What is your job role? | How long have you worked in PERS? Did you have experience of working with OPD population/service prior to being in the PERS? | Can you tell me about what your job within the PERS is? | | What do you do on a day-to-day basis? | How much contact do you have with PERS service users on a weekly basis? |  |
| 3) What factors do staff think are important in the success of PERS offenders? | What do you think contributes to PERS user’s success in open condition? | Do you think the environment of the PERS service user, or the individual characteristics of the of the person are more important for their success? | | How does PERS help service users stop being returned to closed conditions? |  |  |
| *Show the logic model & SR outcomes* |  |  | |  |  |  |
| 4) What do PERS staff think of an I-DAG of progression factors? | What do you think of these diagrams? | Do the diagrams represent your understanding of what may be involved with progression? | | Is there anything you would add to or change about the diagrams? |  |  |
| 5) What do PERS staff think of the logic model? | What do you think of the PERS diagram? | Does the diagram represent your understanding of how PERS works? | | Is there anything in the PERS structure that you would add or change? | Is there anything in the PERS outcomes that you would add or change? | Are there any contextual factors you think are missing? |
| 6) How is PERS being impacted by COVID in both service delivery and outcomes for offenders? | How is the coronavirus outbreak impacting service delivery and service user outcomes? | What parts of PERS have you not been able to deliver due to COVID? | Do you think there will be long-term implications for service users due to coronavirus? | | | |
| 7) Thanks, and closing comments – opportunity to request notes/ transcripts. |  |  |  | | | |

**Supplement 2:**

**Service users interview schedule:**

Questions

Opening Question

How did you get here?*

Follow-up Questions

How has PERS been helpful?

How has PERS not been helpful?

How did COVID-19 impact your time at PERS?

What do you feel is needed for you to succeed?

Did you feel you were in a good place within yourself whilst at the PERS?

How is your mood now?

Community only: How was the transition from open conditions into the community?

*Here will replaced for the situation the service user is in (closed conditions, open conditions, community)
